# Metrics for Accounting Deaths during COVID-19

**DOI:** 10.1101/2020.08.29.20184176

**Authors:** Benjamin Stear, Kyle M. Hernandez, Vidya Manian, Som Dutta, Deanne Taylor, Catharine A. Conley

## Abstract

Since early April 2020, COVID-19 has been the third-largest cause of death in the US, becoming the largest single cause of death during the last two weeks of April. This pandemic has strained public health systems in many states, leading to extended delays in reporting deaths to the US Centers for Disease Control National Center for Health Statistics (NCHS), as well as rumors about the accuracy of reported data. To assess these concerns, we have adapted simple financial statistics to evaluate possible anomalies in mortality data and applied them to NCHS and US Centers for Disease Control Case Surveillance (CS) datasets. NCHS data released 21 October record almost 8000 more deaths from COVID-19 in March and April, US wide, than are captured in the CS daily provisional counts of COVID-19 deaths during those months. For five consecutive weeks during June, July, and August, the states Alabama, California, Florida, Georgia, and Texas each reported at least 40 more deaths per week to the NCHS than were captured in CS daily counts for the same period, with Florida and Texas reporting over 1000 and 4000 more COVID-19 deaths, respectively. In contrast, counties in multiple states attribute to COVID-19 fewer than 10% of deaths reported to the NCHS over the period February-September 2020, while simultaneously reporting over twice as many deaths from all causes compared to the same period in previous years. Metrics based on ratio analysis can provide useful information on the reliability of data used for inputs to epidemiological models.

## Introduction

The COVID-19 pandemic is disrupting nearly all aspects of global society, with widespread health, economic, and sociocultural impacts from both the disease and responses to it. To ensure that pandemic responses are effective but not extreme, it is essential to characterize how the disease is spreading accurately, and as promptly as possible. Predictive models are valuable tools to inform pandemic response, and a large number of COVID-19 models have been developed (1). Unfortunately, published models have not represented US spread accurately, with highly variable outputs between different models, as well as variability between updates to a single model (2).

Regression analysis is a preferred epidemiological approach to model and predict deaths over time, but inconsistencies in the conditions being modeled render regression predictions less reliable, for example when completeness of input data is based on historical values. A caveat often cited when discussing financial scenarios susceptible to anomalous future behavior is “Past performance is no guarantee of future returns” (e.g., 3). Techniques for evaluating financial performance can be applied to death data by treating deaths from individual causes as different accounting streams and evaluating fractions of current-year deaths from all causes as well as ratios with historical data. COVID-19 daily surveillance by the CDC Case Surveillance (CS) Task Force (4) and public pandemic tracking sites provide rapid but possibly inaccurate updates on the numbers of deaths from COVID-19, while the CDC National Center for Health Statistics (NCHS) reports accurate data about deaths including COVID-19, although these data are delayed in time, and may be less than 80% complete even after several months (5).

Here, we analyze data on deaths attributed to a set of ‘select causes’, including weekly deaths from COVID-19 in the entire United States (US) and by state, for consistency over time, and identify indications of potential anomalies. We also analyze cumulative data from the NCHS on deaths from COVID-19 and all causes in all counties for which COVID-19 deaths are reported, compared with data recorded by public pandemic tracking sites including the New York Times (6) and USA Facts (7). Ratio analysis provides novel perspectives on mortality associated with the COVID-19 pandemic.

## Methods

### CDC Datasets

Data on deaths from COVID-19 and All Causes were obtained from the 28 October NCHS dataset “Provisional COVID-19 Death Counts in the United States by County” (8), which provides cumulative data on deaths reported to the NCHS between 1 February and 24 October, 2020, by county. Cumulative counts of deaths from COVID-19 were also retrieved from the New York Times (7) and USAFacts (8) public COVID-19 tracking sites. Data on monthly deaths from All Causes by county from 2016-2018 were retrieved from the NCHS Wonder database (9). Monthly mean and standard deviation values were calculated, and aggregated to generate cumulative average historical data for February-October.

NCHS datasets on “Weekly Counts of Deaths by State and Select Causes”, in final form for 2014-2018 (10) and as provisional data for 2019-2020 (11), were downloaded each week for which updates were made available, since the end of May 2020. These datasets contain weekly counts of deaths as MMWR weeks, starting from the first day of the first year in the dataset, for each of the 50 states, including Puerto Rico and the District of Columbia, with New York City reported separately from New York state. The dataset also reports data for the ‘United States’ that include all 50 states and the District of Columbia, but not Puerto Rico or other outlying US territories. Entries report the number of deaths, aggregated as ‘All Cause’ or ‘Natural Cause’ deaths (‘natural’ means not caused by some external event such as an accident or deliberate action), as well as a set of ‘Select Causes’. The ‘Select Causes’ dataset for 2020 also reports two categories of deaths from COVID-19, as the ‘Underlying Cause’ or from ‘Multiple Causes’. All analyses presented here were performed using only deaths from ‘COVID-19 as an Underlying Cause’.

Daily counts of death from COVID-19 compiled by public tracking sites are very similar to daily counts of death reported by the CDC Case Surveillance Task Force, with only a small number of divergent datapoints, that appear to result from revisions to daily counts made by some trackers and not others (data not shown). As an additional consistency check, weekly counts of COVID-19 deaths were derived in two ways, by summing 7 days of reported daily increases, and by subtracting the previous week values from cumulative counts.

A major objective of the current work is to include all available data in the analyses presented. However, the NCHS suppresses data entries with numbers of deaths between 1 and 9, rendering data incomplete for states with smaller populations and therefore lower weekly counts of death. To perform sensitivity analysis on the lower and upper bounds for missing information, suppressed values were replaced with values between 1 and 9. For the results on ‘Select Causes’ reported here, suppressed values for deaths from COVID-19 were replaced with ‘1’, to take the lower bound on counts of deaths associated with COVID-19, and suppressed values for causes of death other than COVID-19 were replaced by ‘2’, to reduce apparent bias towards COVID-19 in states reporting small numbers of deaths from select causes. No counts were imputed into cumulative county datasets.

The NCHS dataset “Excess Deaths Associated with COVID-19” (12) includes ‘threshold’ values, above which deaths are considered to be excess, and also ‘weighted’ values that give an expected number of deaths predicted by CDC regression algorithms. The ratio of ‘threshold’ to ‘weighted’ values falls approximately 3 standard deviations above the mean of weekly deaths from all causes in 2016-2019 (Fig. 1). Both parameters are considerably less than 10% above the mean historical value for deaths from all causes, and data in 2020 are likely to increase further due to incomplete reporting. Therefore whenever the number of deaths already reported in 2020 is greater than 10% above the historical mean, this number of deaths exceeds both the regression threshold and normal distribution confidence limit.

**Figure 1.**
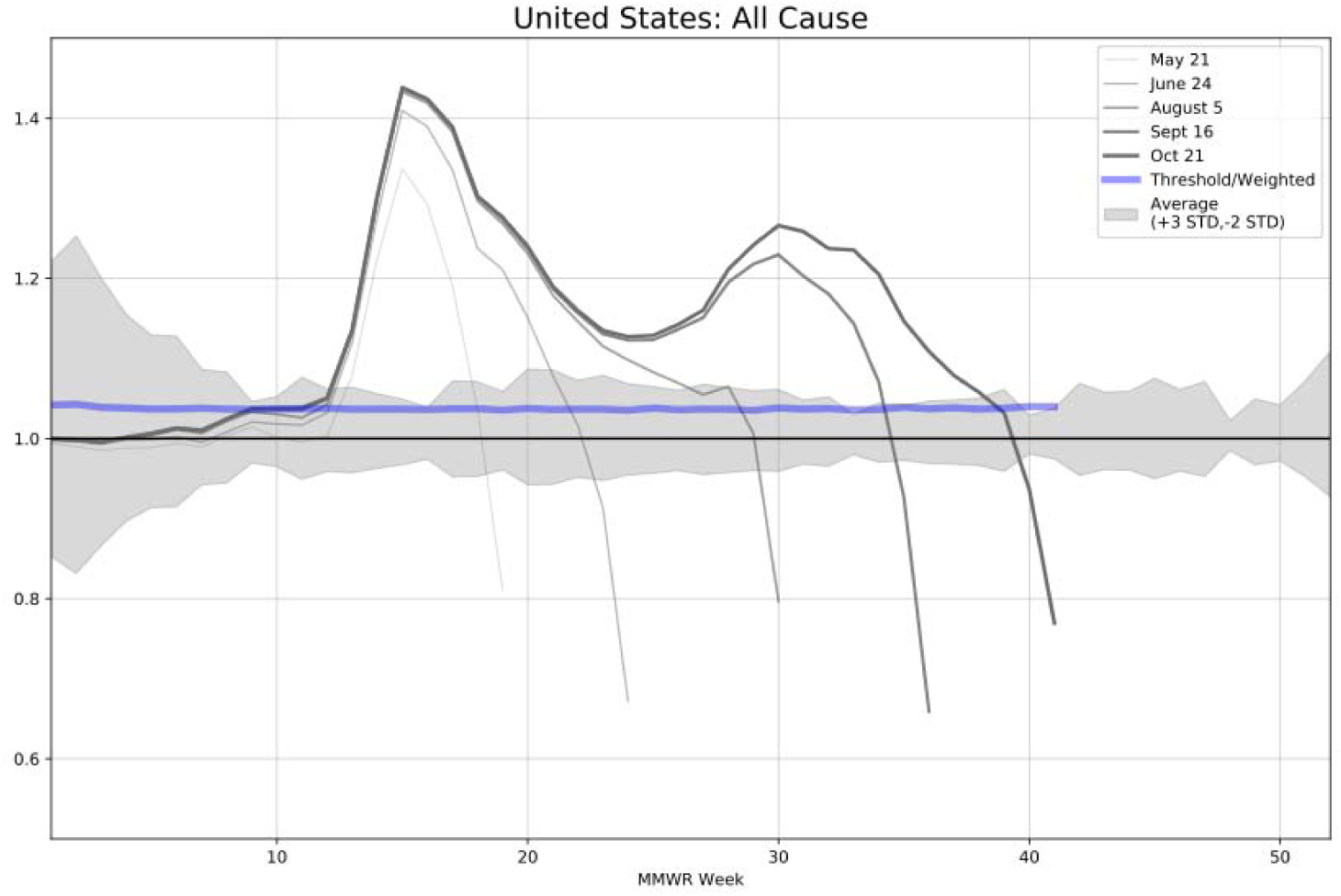
Ratio of 2020:Average(2016-2019) weekly deaths from All Causes in the United States. Traces indicate the ratio of deaths from all causes in 2020 as reported in the NCHS dataset released on the indicated date to the average of deaths from all causes in the corresponding MMWR week for the four years 2016-2019. Grey shading indicates 3 standard deviations above and 2 standard deviations below the mean values: the standard deviations are much greater in the winter due to deaths from prior-year flu epidemics. The purple trace indicates the ratio of the CDC excess death ‘Threshold’ to ‘Weighted’ predictions for deaths from all causes. Between MMWR weeks 10 and 50, values more than 10% away from the mean considerably exceed both threshold estimates and 95% confidence limits for normally distributed data.

Incomplete reporting produces incorrectly low numbers for 2020 data, yet lower populations in prior years would also reduce historical mean counts of death. Although variability in deaths due to influenza greatly increases the standard deviation in historical data for winter months, a limit 2 standard deviations below the 2016-18 mean for cumulative county data over the 9 months February-October also falls well within 10% of the historical mean (Fig. 1).

Considering the multiple sources of error, we refer to data as ‘significant’ when cumulative county deaths from all causes in 2020 are higher than 110% or lower than 90% of historical mean values. Although the exact levels of statistical confidence are uncertain, this approach assures significance of better than 95% confidence.

## Analysis

The programs Orange Data Mining (14) and Microsoft Excel (Microsoft Corp.) were used for data preprocessing and analysis. Single-week averages and standard deviations were calculated individually for each MMWR week across years 2014-2019. Pearson correlation coefficients were calculated for subsets of county data using the ‘Correlations’ widget in Orange. Percentage and ratio calculations were performed independently using both Python and Orange Data Mining. Analyses and supplements are available from the Coronavirus International Research Team Github (https://github.com/COV-IRT/dmwg-covid-reporting-qc/).

## Results

### Weekly Deaths from COVID-19 in the US and Several States

The most basic test for accounting consistency is to compare values for the same accounting stream that are compiled by different entities. In NCHS and daily surveillance data on counts of deaths from COVID-19 (Fig. 2), the numbers of deaths from ‘COVID-19 as an Underlying Cause’ recorded in NCHS datasets for a particular MMWR week increase in subsequent months of dataset releases (Fig. 2, rainbow-colored traces), as expected due to time required for reporting deaths to the NCHS. Daily surveillance counts recorded in the dataset “United States COVID-19 Cases and Deaths by State over Time” (13) compiled by the CDC Case Surveillance Task Force do not appear to be updated after initial release, and the daily death data reported at state level are in general identical to at least one or other of the public trackers, although not always the same public site (data not shown).

**Figure 2.**
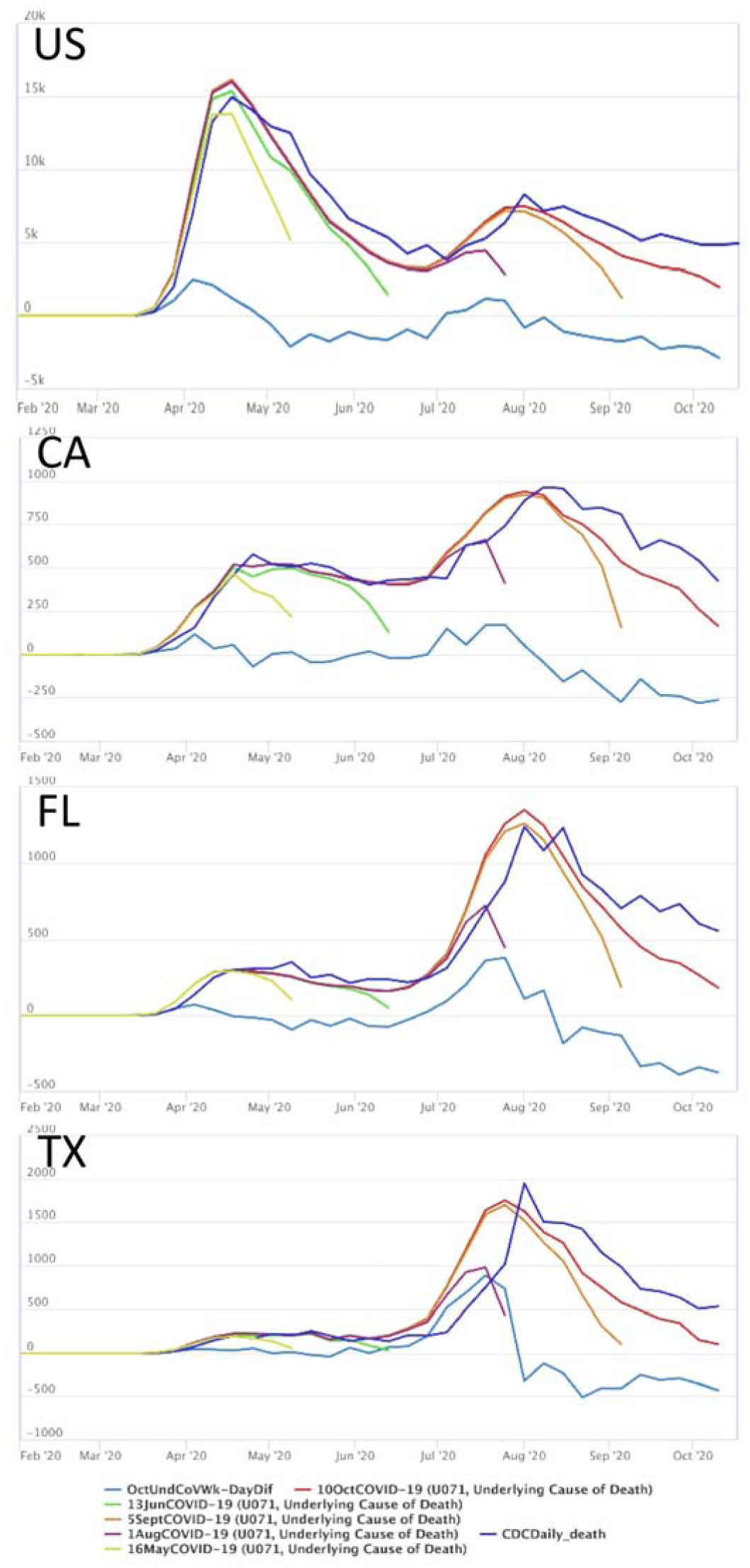
Weekly numbers of deaths from COVID-19 recorded in CDC daily surveillance and NCHS datasets. Trace colors indicate date of most recent weekly data: 16 May is yellow, 13 June is green, 1 August is purple, 5 September is orange, and 10 October is red. Dark blue trace indicates CDC daily surveillance counts aggregated by week. Light blue trace indicates the difference between the 10 Oct NCHS dataset and surveillance data.

Comparing five months of NCHS datasets with weekly data on COVID-19 deaths aggregated from the 30 October release of daily surveillance data (Fig. 2, blue traces) identifies inconsistencies between the two datasets. For weekly deaths in the US as a whole (Fig. 2, US), NCHS datasets released after June record weekly numbers of COVID-19 deaths in March and April that exceed the CS surveillance data on COVID deaths in the same weeks. NCHS datasets released in September and October record higher numbers of COVID-19 deaths also for weeks in July, with additional deaths from COVID-19 in July and August still being reported in NCHS datasets released as of late October.

California, Texas, and Florida are the three most populous US states. California (Fig. 2, CA) experienced an early onset of COVID-19, yet the difference between daily surveillance and NCHS death counts never exceeded 250 per week, with the greatest difference detectable in July. Florida (Fig. 2, FL) experienced a smaller outbreak than California early in the pandemic, yet in July several hundred more COVID-19 deaths per week were reported in the October 21 NCHS dataset, than were recorded by daily surveillance data: over 1000 deaths in total. Prior to June, Texas (Fig. 2, TX) experienced the smallest outbreak of the three states, but in late June and July more than half the COVID-19 deaths, as reported for Texas in the October 21 NCHS dataset, were not recorded in CS daily surveillance data (light blue trace higher than dark blue). Cumulatively, between 15 June and 20 July, for Texas the NCHS reports over 4000 COVID-19 deaths that were not identified in the daily surveillance data.

### Cumulative Deaths from COVID-19 by County

Comparing weekly data gives snapshots in time, but does not provide comprehensive information regarding historical trends, nor permit comparison across accounts of different sizes. In finance, comparing current-period with historical data is called ‘horizontal’ analysis, while normalizing accounts of different sizes by considering individual accounting streams as fractions of a relevant total is called ‘common size’ or ‘vertical’ analysis (3). We performed both types of ratio analysis using 2020 data on cumulative deaths from COVID-19 and All Causes from public tracking sites and the 28 October NCHS dataset on “Provisional COVID-19 Death Counts in the United States by County”, compared with historical data on county-level deaths from All Causes in 2016-2018 over the period February-October.

As noted above, an important consideration when comparing deaths in 2020 to historical data is that data from 2020 are incomplete, so cumulative death counts for 2020 will almost certainly increase. Despite this probable incomplete reporting, a substantial number of counties have already reported significantly more deaths from all causes in 2020, relative to the same county’s historical 2016-2018 mean values (Fig. 3A). Plotting county-level data by state and region facilitates visualization of differences in geographic distribution: counties from states in the South (S) and Central (C) regions of the United States more often report ratios of deaths in 2020 that are more than two times the 2016-2018 average.

**Figure 3.**
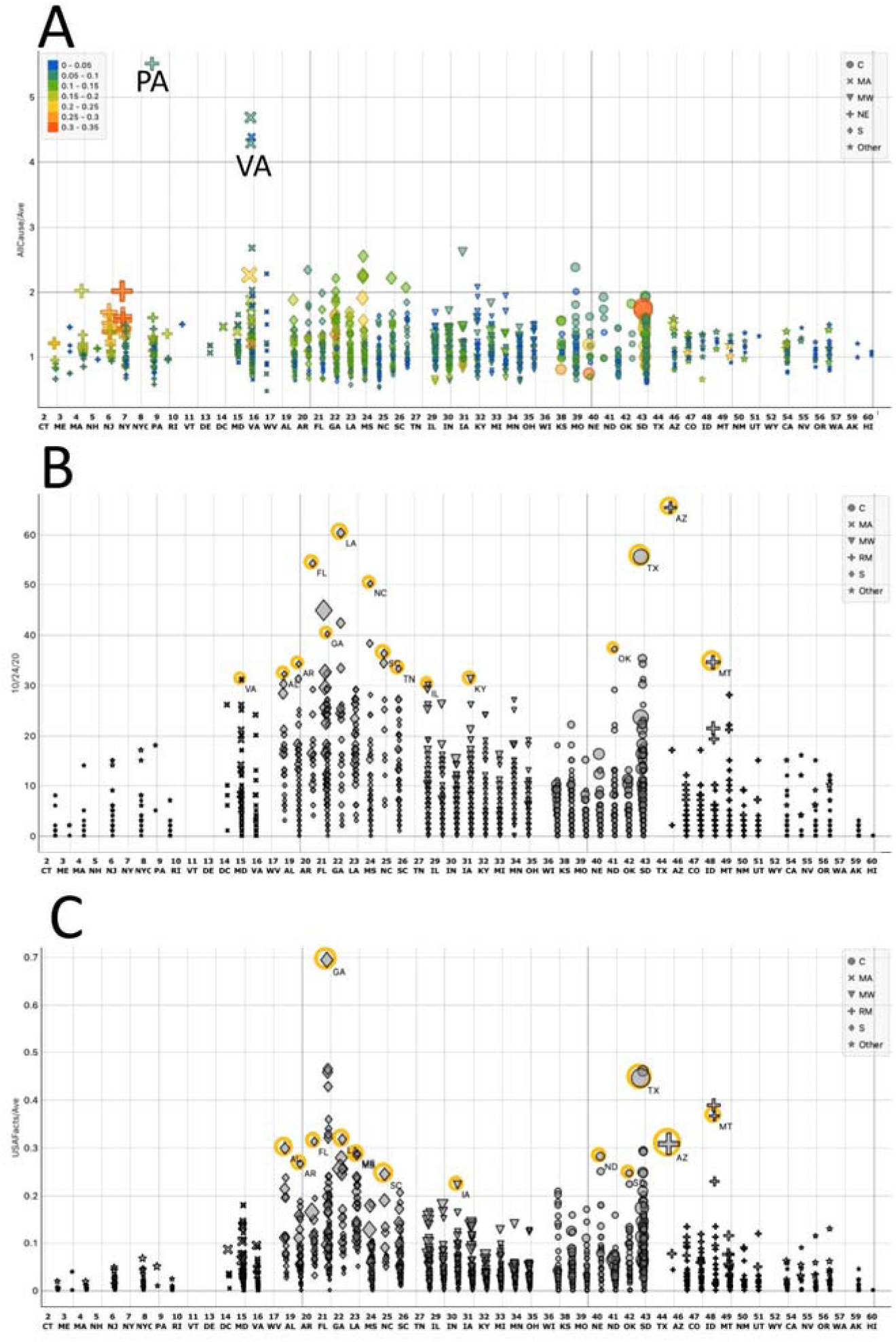
Cumulative deaths from COVID-19 by County. A) NCHS-recorded deaths from All Causes divided by the average number of deaths for February-October in 2016-2018. Color indicates percentage of deaths from COVID-19: size proportional to the ratio of deaths from COVID-19 to 2016-2018 average deaths from all causes. B) Number of deaths captured as cumulative counts of COVID-19 deaths by USAFacts, in counties for which the NCHS has recorded no deaths from COVID-19. Size proportional to the the ratio of deaths from COVID-19 over the 2016-2018 average of deaths from all causes. C) Ratio of COVID-19 deaths recorded by USAFacts but not the NCHS, to average deaths from all causes; size proportional to the number of COVID-19 deaths recorded by USA Facts.

A further challenge to performing comprehensive analyses is the extent of missing data: the NCHS does not record deaths from COVID-19 in 1582 counties, almost half the counties in the US across nearly all states, in data through 24 October (Fig. 3B). In these same counties, the public tracking sites recorded more than 12,000 COVID-19 deaths : 12,066 total deaths counted by USA Facts and 12,171 counted by the New York Times. For multiple counties in Georgia, as well as several counties in Texas and Montana, the number of deaths counted by public tracking sites exceeds 30% of the average deaths from all causes in 2016-2018 (Fig. 3C), although the NCHS has not yet recorded any deaths from COVID-10 in these counties.

Considering only the half of US counties for which the NCHS has recorded deaths from COVID-19 (Fig. 4), further anomalies are revealed by examining correlations between cumulative (February-October) percentages of deaths from COVID-19, with the ratio of deaths from all causes in 2020 to 2016-2019 averages. Many counties, from nearly all states, have reported deaths from All Causes in February-October 2020 that are over 10% higher than the 2016-2018 average deaths from the same period (Fig. 4A). Counties in PA, VA, WV, AR, IA, KY, and MO have reported more than twice as many deaths as average to the NCHS; however at the same time fewer than 10% of these deaths are recorded in the NCHS dataset as being due to COVID-19 (Fig. 4A, blue shades). Conversely, a number of counties, though from fewer states, have reported less than 90% of the average number of deaths from all causes for February-October (Fig. 4B). Among these, counties from VA, AL, AR, FL, LA, MS, IN, IL, MN, NE, TX, and ID, have reported fewer than three quarters of the average number of deaths from all causes, yet at least 10% of the deaths that have been reported are due to COVID-19 (Fig. 4B, tan shades). The most extreme outlier is Perkins County, Nebraska, where 34% of deaths reported in 2020 are due to COVID-19, although less than 72% of the average number of deaths have been reported.

**Figure 4.**
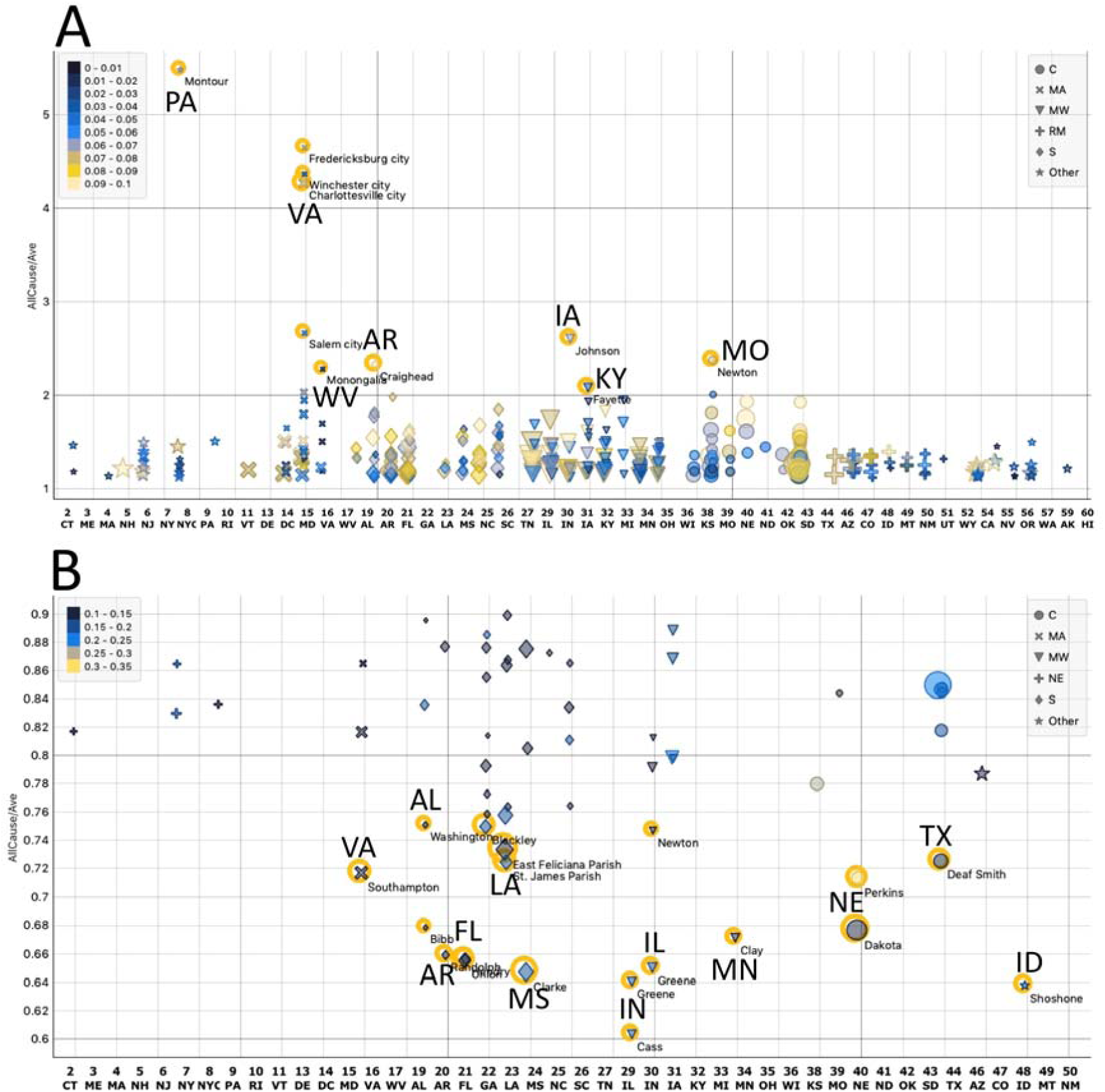
County level cumulative deaths from COVID-19 recorded by the NCHS between 1 February and 24 October. Each point represents a single county, with color indicating the percent of deaths from COVID-19 as an Underlying Cause recorded by the NCHS, and size representing the relative percentage of deaths from COVID-19 recorded by USAFacts. A) Counties with NCHS data indicating that deaths from all causes exceed 110% of 2016-2018 historical mean values, and also less than 10% of these deaths are reported as due to COVID-19. Labelled points indicate states with at least one county reporting at least twice the average number of deaths from all causes, yet at the same time report fewer than 10 percent of these deaths as due to COVID-19. B) Counties with NCHS data indicating that reported deaths from all causes in 2020 are less than 90% of 2016-2018 historical mean values, yet at the same time more than 10% of reported deaths are due to COVID-19. Labels indicate states with at least one county reporting under three quarters (75%) of historical mean deaths, with over 10% of these deaths from COVID-19.

Across all counties reporting fewer than 90% of average deaths and also at least 10% of deaths due to COVID-19, the Pearson correlation coefficient for NCHS data is 0.18. Pearson correlation coefficients for correlations of the ratio of NCHS data on 2020/average deaths from all causes with the percentage of COVID-19 deaths using data from daily trackers are actually negative, at - 0.24 for New York Times data and −0.22 for USAFacts.

## Discussion

The US Dept. of Health and Human Services’ *COVID National Diagnostic Strategy* (15) recommends using deaths from COVID-19 as reported to the CDC as one criterion to inform policy decisions on reopening. A number of online tools evaluating the current status of reopening readiness also use CDC-reported death data as inputs (e.g., 16). Evaluating the completeness of reported deaths from COVID-19 is an important element of strategies for ensuring that the US reopens safely. Further, assessing the extent of possible under-reporting is a key aspect of understanding the accuracy of death data for use in epidemiological models.

Unfortunately, analyses are only as good as the data used to make them, and the data available on deaths from COVID-19 in the US are highly unreliable. The CDC notes that death data are only 75% complete after 8 weeks under normal circumstances (5), and delays during the COVID-19 pandemic have rendered data even less complete than normal (12). Early in the pandemic there was considerable confusion about attributing deaths to COVID-19, but deaths should have been reported more consistently after CDC guidance on counting deaths from COVID-19 was released on 14 April (17). Even so, the CDC daily surveillance data documenting deaths from COVID-19 do not capture correctly all deaths from COVID-19 that are reported to the NCHS, with some states showing much larger discrepancies than others (Fig. 2).

In 2020, the US as a whole has reported significantly greater than average numbers of deaths from all causes (Fig. 1). However, a considerable number of counties have reported significantly fewer deaths than average (Fig. 3), and some counties reporting significantly fewer deaths than average simultaneously report greater than 10% of these deaths as due to COVID-19 (Fig. 4). In counties across the US, where reporting of deaths from all causes is more noticeably delayed, it appears that larger percentages of deaths from COVID-19 are also being captured by public COVID trackers. The negative correlation of COVID-19 data recorded by public tracking sites with NCHS data on deaths from all causes elevates concerns that extended delays in reporting could have a considerable impact on the accuracy of deaths counts recorded by the CDC, and by extension US understanding of the severity of -- and effectiveness in responding to -- the COVID-19 pandemic.

As COVID-19 continues to spread during the coming winter, from a public health perspective it will be important to address inconsistencies between daily surveillance data and deaths eventually reported to the NCHS.

## Data Availability

Data cited in the MS are public at the cited links. Analytical code is available at the cited links.

https://github.com/COV-IRT/dmwg-covid-reporting-qc/

## Acknowledgments

We thank the founders and members of the COVID-19 International Research Team, Dr.s Afshin Beheshti, Todd Treagan, and Krista Ternus, for facilitating this collaboration; and particularly members of the COV-IRT Modeling Subgroup, particularly Dr.s Daniel Guimarães Tiezzi, Sylvain Costes, Talayeh Razzaghi, and Mohammed Eslami, for critical discussion of the analyses.

## References

[1] J. T. Unwin et al., “Report 23: State-level tracking of COVID-19 in the United States,” no. May, 2020. doi: https://doi.org/10.25561/79231

[2] I. Holmdahl and C. Buckee, “Wrong but Useful — What Covid-19 Epidemiologic Models Can and Cannot Tell Us.”

[3] Investopia, “Financial Analysis.” [Online]. Available: https://www.investopedia.com/financial-analysis-4427788

[4] Centers for Disease Control “https://data.cdc.gov/Case-Surveillance/United-States-COVID-19-Cases-and-Deaths-by-State-o/9mfq-cb36”.

[5] National Center for Health Statistics “Technical Notes on Provisional Death Counts for Coronavirus Disease (COVID-19).” [Online]. Available: https://www.cdc.gov/nchs/nvss/vsrr/covid19/tech_notes.htm.

[6] New York Times https://github.com/nytimes/covid-19-data.

[7] USA Facts https://usafacts.org/visualizations/coronavirus-covid-19-spread-map/.

[8] National Center for Health Statistics “Provisional COVID 19 Death Counts in the United States by County.” [Online]. Available: https://data.cdc.gov/NCHS/Provisional-COVID-19-Death-Counts-in-the-United-St/kn79-hsxy

[9] Centers for Disease Control “Wonder database.” [Online]. Available: https://wonder.cdc.gov/.

[10] National Center for Health Statistics “Weekly Counts of Deaths by State and Select Causes, 2014-2018” [Online]. Available: https://data.cdc.gov/NCHS/Weekly-Counts-of-Deaths-by-State-and-Select-Causes/3yf8-kanr.

[11] National Center for Health Statistics “Weekly Counts of Deaths by State and Select Causes, 2019-2020.” [Online]. Available: https://data.cdc.gov/NCHS/Weekly-Counts-of-Deaths-by-State-and-Select-Causes/muzy-jte6.

[12] National Center for Health Statistics “Excess deaths associated with COVID-19.” [Online]. Available: https://www.cdc.gov/nchs/nvss/vsrr/covid19/excess_deaths.htm.

[13] Centers for Disease Control “United States COVID-19 Cases and Deaths by State over Time.” [Online]. Available: https://data.cdc.gov/Case-Surveillance/United-States-COVID-19-Cases-and-Deaths-by-State-o/9mfq-cb36

[14] Demsar J, Curk T, Erjavec A, Gorup C, Hocevar T, Milutinovic M, Mozina M, Polajnar M, Toplak M, Staric A, Stajdohar M, Umek L, Zagar L, Zbontar J, Zitnik M, Zupan B (2013) Orange: Data Mining Toolbox in Python, Journal of Machine Learning Research 14(Aug): 2349–2353.

[15] Dept. of Health and Human Services, “COVID National Diagnostic Strategy,” 2020. [Online]. Available: https://www.democrats.senate.gov/imo/media/doc/COVID%20National%20Diagnostics%20Strategy%2005%2024%202020%20v%20FINAL.pdf

[16] “Covid Exit Strategy.” [Online]. Available: www.covidexitstrategy.org.

[17] Centers for Disease Control “About CDC COVID-19 Data” [Online]. Available: https://www.cdc.gov/coronavirus/2019-ncov/cases-updates/about-us-cases-deaths.html

